# A Holistic Ecosystem for Clinical Protocols and Trials Management

**DOI:** 10.1101/2024.03.10.24304032

**Authors:** Stella C. Christopoulou

## Abstract

The proposed Holistic Ecosystem for clinical Protocols and Trials Management (clinicalNET) project aims to establish a human and technical network of healthcare researchers, business partners, and stakeholders. The consortium will include specialists from various fields, including healthcare management, medicine, computer engineering, marketing, publishing, and public relations. The project will design and implement a semantically enabled information system for representing data and knowledge gathered during clinical trials. The clinicalNET project will define specifications for healthcare management services and knowledge engineering techniques, creating an intelligent collaborative web tool for information production and provision in healthcare. It will follow EU health policies and promote participation from leading researchers, stakeholders, and organizations. The clinicalNET will provide a health holistic ecosystem approach with open standards, trusted networks, and co-creator networks.

## 1. INTRODUCTION

The objective of the proposed Holistic Ecosystem for clinical Protocols and Trials Management (clinicalNET) project is the creation of a human plus technical network composed of researchers, business partners and stakeholders in the field of healthcare. The consortium will include specialists in the fields of healthcare management, medicine, computer engineering, marketing, publishing, and public relations. Therefore, the context of the proposal is two-fold. Firstly, it will focus on designing and implementing a semantically enabled information system that will allow for the representation of data and knowledge gathered during the conduct of clinical trials. Thus, an information map according to existing or new knowledge bases will be developed, that will allow for reusability of data under different points of view. Existing efforts on the field attempt to semantically model only the metadata of a clinical trial. We envision the development of a system that will be able to include eligibility criteria as well as results.

Secondly, as a development of an earlier premature work [1] the proposed project will define the specifications of healthcare management services and knowledge engineering techniques towards an intelligent collaborative, integrated and participatory web tool for production and provision of information, new knowledge, and services in healthcare. It will act as a common place of communication among stakeholders in the field of clinical protocols and trials, information technology etc., but patients too.

As a result, a multi-discipline human network will be developed which will be able to communicate and compile scientific progress for the benefit of patients and the society.

### 1.1. Mission and Policies

Clinical trials are a precise useful tool for the development of medical research and the application of medical and healthcare and the further utilization of medical knowledge in the development of clinical guidelines and the provision of personalized health services. Thus, in the clinicalNET is proposed a fundamentally different and decentralized approach and a wiki-semantic Web tool for clinical protocols and trials management. In the proposed *clinicalNET*, the management of the clinical trials at all stages is a co-working process and is supported by novel technological methods in order to ensure cooperation, exchange of appropriate medical information, possibility of extraction of medical knowledge and its use in an accurate, timely and complete manner.

The means for the implementation of the *clinicalNET* are the following:

○ The researchers, policy makers, professionals, organizations, etc.
○ The technological Platform that supports the Worldwide cooperation and inclusiveness of researchers, standardization, and re-use of information.
○ Innovative and effective policies. On the one hand, the *clinicalNET* will follow the EU health policies, but on the other hand will establish a plan towards revising them on a basis of innovation and effectiveness.

The main procedures of this plan will ensure the following:

○ Promotion of the participation in the activities of the *clinicalNET* leading researchers, stakeholders, organizations etc. from participating countries in order to ensure the maximum possible development and dissemination of knowledge and applications in the field of *clinicalNET* strategy is to attract and cooperate with top-scientists based on three pillars: Firstly, detailed information about the *clinicalNET* will be made available via the *clinicalNET’s* website, newsletters, videos, posters, presence at social media, workshops and other dissemination methods. Secondly via inviting researchers that have a strong academic profile. Fit for Health 2.0 etc. and at conferences, publications, workshops. Finally, researchers will be attracted via the dissemination and exploitation activities of the *clinicalNET* participants.
○ All activities of the *clinicalNET* will be shared in as many as possible participating countries, i.e. will be given leadership roles at *clinicalNET* level in different countries and conducting *clinicalNET* meetings, workshops and conferences in different ways each time participating Universities/Institutes/Organizations.

Thus, the *clinicalNET* will achieve its mission which is to provide a health holistic ecosystem approach that has the standards of an open, constantly evolving, as most as possible trusted, independent, co-working and co-creator NET in providing healthcare information and management in evidence-based practice, and further support in research and practice for researchers, organizations and individuals.

## 2. THE HOLISTIC ECOSYSTEM FOR CLINICAL PROTOCOLS AND TRIALS MANAGEMENT (clinicalNET)

### 2.1. Description of the proposed Project

Despite the growing interest and the challenges associated with collaboration in the process of development and adaptation of knowledge in medical practice on the web there is a lack of a common methodological framework that will put together relevant experiences, tools and information in the field. Clinical protocols and trials is a challenging area because healthcare professionals must manage, collaborate and share a large amount of information from different sources and work together to acquire knowledge [2].

The *clinicalNET* is the coordination of research efforts to develop an interdisciplinary Worldwide net for collaborative, context-aware management and exploitation of clinical protocols and trials. It will also focus on the development of an adequate management framework through an open (wiki) and semantic web 3.0 platform as an acceptable, feasible and effective solution.

### 2.2. Relevance and timeliness

Innovation in medical knowledge management is the starting point for *clinicalNET*. Clinical protocols and trials come in different formats and no predefined structure or metadata is identified. There is a need to identify and transcript clinical protocols and trial’s structure, provide access to medical knowledge, strengthen collaboration among multidisciplinary stakeholders and investigate the development of a business model adequate to the health sector needs for resource conservation without undermining but improving the Quality of Life (QoL) that patients experience.

### 2.3. Research Coordination Objectives

While vast research and industry efforts focus on distinct disciplines in the field, there is currently no systematic application of an interdisciplinary methodology that will merge knowledge, experiences, and tools.

A common framework would enhance not only medical knowledge dissemination and application but collaboration and context-aware management of clinical protocols and trials too. Towards this, *clinicalNET* will offer:

⍰ Coordination of research on modeling and management of clinical protocols and trials
⍰ Exploitation of relevant research and value-added business activities.
⍰ The creation of a Global network of health-care stakeholders (e.g. professionals, clinical researchers, nurses, health-care managers and policy makers, health information specialists) for the development, evaluation and dissemination of “best practices” on clinical protocols and trials management.
⍰ Dissemination of research outcomes in the field of evidence based medical practice by employing modern technologies such as open and linked data, semantic and wiki techniques.
⍰ The design, development, and management of an open and semantic platform for clinical protocols and trials management. The potential users of the platform will be trained and will evaluate it.
⍰ Support cooperation between health professionals and organizations to form a networked community of experts.

## 3. THE INNOVATIONS THAT INTRODUCES THE ClinicalNET

### 3.1. Description of the state-of-the-art

As medical information increases very rapidly, practitioners must stay updated with relevant literature and research.

Clinical protocols and trials are constantly increasing, and safety directives demand larger patient participation. Increased patient enrollment in trials is crucial for effectiveness, efficiency, and safety in healthcare. Interest in best clinical practices, evidence-based medicine [2] and data quality integration is growing. Domain knowledge must be refined while sophisticated data analysis can help deduce common or supplementary results from diverse clinical trials. Clinical trial taxonomy can be based on observational or experimental studies [2].

### 3.2. Innovation in tackling the challenge

The healthcare domain is crucial for improving life quality, but medical data and knowledge flow between different systems are inefficient. A proposed system, aims to overcome automation deficiencies in medical data monitoring

Contemporary applications propose the implementation of many different technologies and models like dedicated ontology, semantic or wikis schemas, multi-agent systems [3] to fill the gap between healthcare systems and care provision communities, blending next-generation distributed services in healthcare [4].

More specifically, the main challenges that face the *clinicalNET* and the corresponding innovations that this proposes in order to manage them effectively are the following:

⍰ *Challenge: Huge and constantly growing volume of information*
⍰ *Innovation: Implementation of an open web platform of clinical trials management systems that support co-working and co-creating by a multidisciplinary research group management model*.

The rapidly and constantly growing volume of information resulting from clinical trials requires state-of-the-art solutions to manage them. The proposed solution includes both the application of new technological tools (i.e. semantic, wiki approaches) and methodologies (i.e. application of co-working and co-creating models). All these tools and methods are the first to be studied, tested and evaluated in the domain of clinical trials management.

Moreover, this *clinicalNET* will be constantly supported by collaborating multidisciplinary research groups that will focus on the finest models and motivate the applications of best practices and services by the industry.

This proposes a new holistic concept in medical practice and more specifically in clinical trials management. A new model will be proposed which will tackle the above-mentioned challenges as a constantly evolving workflow system, incorporating collaborative methods and innovative technological tools. Furthermore, it will incorporate pre-clinical and clinical protocols and adapt them as an interconnected grid of tasks and resulting information. The management of pre-clinical and clinical protocols will promote:

○ Suitable clinical protocols and clinical trials development.
○ Sponsor tracking for the conduct of the clinical trial.
○ Partnerships established for the conduct of the clinical trial.

Patient recruitment for the successful conduct of the clinical trial. Relevant security and privacy issues will be fully ensured.

Thus, *clinicalNET* will implement a Web 3.0 platform which will offer to scientists and companies novel business models for the management of provided information. Moreover, except the widespread dissemination that will be ensured by a continuous and broad synergy of an interdisciplinary scientific human network via the procedures of *clinicalNET* what is also currently required is to adapt these features to the increased demands by scientists in health and healthcare delivery and especially in terms of reliability and safety. Therefore, in the *clinicalNET* the scientific human network will collaborate on developing a web platform that will support the:

○ development and adaptation of the appropriate applications,
○ successful and fast penetration and utilization of these proposed technological solutions in medicine and health,
○ study and implementation of rules, regulations, ethics and safety at EU and country level.

⍰ *Challenge: Reliability and provision of innovative internet services*
⍰ *Innovation: Development and application of novel business models in conjunction with web 2*.*0 & 3*.*0 technologies* An extremely important issue over the internet is information reliability. This issue is even more crucial when this information relates to patients’ medical profile or public health. Additionally, the stakeholders’ requirements for support and delivery of new or personalized health services are constantly increasing. Therefore, researchers and industry in the field must collaborate towards joint business models that will be assessed by multi-disciplinary consortia as the one proposed in the *clinicalNET*.

## 4. RESOURCES REQUIRED

Capacity building in the scope of the *clinicalNET* focuses on the development of both human and organizational skills in Global level. Participation support, knowledge exchange and processes reform are the main cornerstones towards capacity building. More specifically the activities that have to be adopted include:

○ Partnership, cooperation with and engagement of stakeholders.
○ Training support of participating Institutions/Organizations/ businesses.
○ Organization of workshops at the participating countries tightly connected with the outcomes of the *clinicalNET*.
○ Joint training and interaction among governmental officers, business partners, researchers, students and volunteers.
○ Joint planning process aligned to country processes and needs towards priority setting of beneficiary countries.
○ Shifting abilities for management of clinical trials, access to knowledge and collaboration in healthcare to individuals and non-profit organizations.
○ Transition of responsibilities to individuals.
○ Promotion of scientific connections that will bridge gaps in know-how and support continuous development of the domain.

### 4.1. Technological needed resources

A wiki can be identified not only as a collaboration platform but a potentially enriching learning and teaching tool, that is easy to use, offering rapid deployment, and facilitation of knowledge transfer. Thus, wiki based collaboration systems have been proposed as a solution to promote cooperation between stakeholders in the healthcare sector [2].

Moreover, semantic wikis have been applied for collaborative authoring of biomedical vocabularies supporting patients’ daily life and clinical protocols modeling.

At the same time, considerable research efforts have focused on the task of formalizing clinical trials and their eligibility criteria. Many researchers introduce a patient-oriented semantic integration approach and semantically linked clinical research ontologies is proposed. Also, there is a continuously developing research in the field of the automatic semantic annotation methods and search of a target corpus using several knowledge resources. More specifically in the biomedical domain, there are several clinical text analyses, knowledge extraction and annotation systems that map free biomedical text to standard conceptual form using metathesaurus.

The access and the exploitation of the *clinicalNET* data will be defined by the overall project development team.

## 5. DISCUSSION

The innovative features and benefits of the proposed *NET platform* are the following:

○ Collaborative and perpetually enriched open web platform (wiki) with clinical data management services.
○ Data exploitation, by adding semantic context. Scalability, evolution and collaboration.
○ The functionality of the open and semantic web platform is summarized below:
○ The user will be able to manage its own profile or data with appropriate access rights, to link them with other semantic data and to restructure, enrich and share new information and knowledge generated after processing via relevant tools such as semantic annotators.
○ The system will support online collaboration and co-creation of medical knowledge between users with the same scientific and research interests.

Finally, as the issues of confidence and quality of medical information are identified as of critical importance during their life cycle, this platform will not allow anonymous data modification.

Access to medical content follows the approach already proposed in the previous related work. In that proposed health ecosystem the Users/Groups management module enables users to access functionality and data in order to collaborate and co-create health care content while protecting medical information. Users, user-groups and access rights are deployed so as to implement a security model that protects against inappropriate or non-authorized content publication.

### 5.1. Added value of networking

The proposed platform will support Semantic Web technology. Evidently Web 3.0 Semantic Web technologies (e.g. RDF, OWL, SKOS, SPARQL, etc.) and Linked Data offer added value to information on the web. Open and linked data can be further enhanced through their connection with other data or mining of knowledge. A relevant example is the ability offered to an individual to trace the most appropriate treatment for a specific patient profile. Another example is the ability for a physician, clinician, or organization to find ongoing or recently completed clinical trials for a specific disease area. The development and tracking of these technologies in conjunction with the provided services to the medical researchers are an important issue of high added value.

However, an essential point for the growth of a critical mass for the acceptance and widespread use of these technologies in the health sector will be their ease of use and the provision of training to key-users.

Moreover, nowadays the need for personalized and reliable patient support is widespread. Thus, an added value of the *clinicalNET* is the development of novel health tools e.g. personalized healthcare delivery systems such as they arise as added value of knowledge. Until now, the design and development of modern information systems usually start from scratch. This could be overcome if the existing information and knowledge on the web becomes available to information systems as “re-usable information or knowledge”. For example, the implementation of an home-based information system for a diabetic’s monitoring and consultancy services using biosensors, wireless and mobile services could embed the appropriate for this case clinical guideline which would result as an added value service from the clinical trials management web platform.

Also, from the perspective of addressing the need for searching, retrieving and linking data that exist anywhere on the internet in order to provide integrated information or knowledge, the proposed platform is used as a data integration system that interlinks any heterogeneous data sources and integrates them into global Linked Open Data space. In this case the key value is not the actual information but its potential for association with other information available on the internet (e.g. the correlation of the fundamental data of a clinical trial with the results of the clinical trial). So, in the proposed web platform, the provided data are active links because the provided information is constantly growing and supported with easy and semi-automatic / automatic modes and embedded infrastructures.

Specific consortia, derived and produced from the initial NET of the *clinicalNET* may create and perpetually manage the data using an appropriate tool that will operate as an editing solution and a value-added mechanism for distributed and syndicated structured semantic content on the World Wide Web.

### 5.2. Progress beyond the state-of-the-art

Evidence-based medicine (EBM) is the process of systematically reviewing, appraising, and using clinical research findings to aid the delivery of optimum clinical care to patients [5]. The Cochrane Library [6] with the objective to facilitate the preparation of systematic reviews of randomized controlled trials of healthcare, contains high-quality, independent evidence to inform healthcare decision-making [7].

But, towards the next generation of healthcare services based on new technological tools and structures, evidence-based medicine and consequently the management of clinical trials emerges as a critical issue and requires a novel approach and a more integrated treatment. Clinical trials are a precise and useful tool for the development of medical research and the utilization of medical knowledge in the evolution of clinical guidelines and the provision of personalized health services.

Thus, *clinicalNET* proposes a fundamentally different and decentralized approach and an innovative management tool for clinical protocols and trials. In the *clinicalNET*, the management of the clinical trials at all stages is a co-working process and is supported be novel technological methods (wiki and semantic technologies) in order to ensure cooperation, exchange of appropriate medical information, possibility of extraction of medical knowledge and its use of an accurate, timely and complete manner.

Moreover, through the prism of the dynamically increasing medical information and the necessity for knowledge classification and dissemination to the stakeholders, the clinicalNET will aim at providing a wiki based and semantically enabled platform for clinical protocols and trials management. The offered services will include functionality that is of industry and medical interest:

○ *Patient recruitment:* Focuses on applying the necessary techniques in finding the qualified patients for a trial.
○ *Trial suitability:* Checks whether a clinical trial’s criteria make it appropriate for a specific patient.
○ *Data usability:* Verifies that the clinical trial outcomes can be merged, aggregated or integrated in order to accomplish systematic reviews or unified results presentation.
○ *Health Professional enrolment:* Facilitates the collaboration among stakeholders in terms of access authorization policies on content.

### 5.3. In relation to existing efforts at European and/or international level

A scientific, open, and continuously expanding human NET that may develop and maintain a context-aware and decentralized management platform will foster reusability and dynamic knowledge management. The decentralized management of information deriving from clinical trials is proposed by this clinicalNET, as the best solution for the management of huge and constantly growing volume of data due to the nature of the clinical trials data, and the constant development of new clinical trials. However, the major priority is the determination of the policies to ensure the reliability of these data.

Until now Wikipedia () is the dominating example. The applied cooperation policies and guidelines are proven extremely successful since the provided data are extremely reliable and errors or misleading information is usually corrected real-time.

*LinkedCT* was the first project in the field of semantic representation of medical knowledge that publishes on the web clinical trials as linked data improving their adaptability and usability. A database is formed by converting data sources in RDF and identifying semantic links between data [8], [9].

Moreover, a context-aware approach has been developed in the PONTE project, for effectively guiding medical researchers during clinical trial protocol design and allowing for more efficient and effective access to scientific literature. The suggested approach incorporates intelligent services and advanced text mining mechanisms for scientific literature querying and mining during protocol design, considering the study context (i.e. active substance, target and disease) and the domain context in literature [10].

But, as we mentioned above is the first time, to existing knowledge, that will be studied, implemented, and evaluated in the domain of clinical trials management a real-time co-working and co-creating method as it is possible via the implementation of a wiki and semantic platform. Furthermore the whole project will be supported by and for an open scientific community and the markets (creating a perpetual cycle of processes) via dissemination procedures (workshops, papers, manuals, white papers, etc.).

### 5.4. Provision of qualitative and reusable information. – Innovation: Standardization of information

If the medical information does not strictly obey medical standards (e.g. SNOMED CT, Mesh, ICD-11 etc.) and rules it becomes useless. Standardization of information provides a common understanding/context to anyone (i.e. information systems and human experts) and makes it possible to reuse. Also, standardization rules benefit the rapid development of new information and its management with more affordable resources. Therefore, the standardization of medical information and knowledge related to clinical protocols and trials becomes a necessity.

In parallel, due to the evolution of ICTs associated with improved communications and the management of information and knowledge (Big Data, Data Science), there is a revolution in the concepts associated with the Clinical and Trials management protocols. This is because the healthcare information systems have a large amount of information which is suitable for use in clinical research protocols and the reverse. This revolution in concepts must be managed properly; otherwise there will be a lot of lost resources.

More specifically, a problem that is being addressed, with a great effort by health systems at regional, national, and European and Global level is sustainability, due to demographic changes. The approach is to develop research and development of new technological solutions to favor the coordination of health systems and social services to promote continuity of care in an efficient, effective, quality and sustainable manner.

Another approach is to give more information and knowledge to the patient and their caregivers, to be active actors in the care team. This is also posing new challenges that need answers with the development of new paradigms that involve changes in the development of clinical protocols.

In this *clinicalNET* will be studied the feasibility of developing a standard for Clinical Protocols and trials Management, similar to the ISO 13940 standard “Health Informatics - System of concepts to support continuity of care”, the standard purpose of this is to define the generic concepts needed to achieve continuity of care. The overall aim standard for this is to provide a comprehensive, conceptual basis for content and context in healthcare services. It should be the foundation for interoperability at all levels in Healthcare Organizations and for development of information systems in healthcare [11]. But the healthcare research processes are not in the scope of this standard. There is a lack that will be treated and will be managed in the clinicalNET [12], [13], [14].

### 5.5. Expected Impacts

The *clinicalNET* project aims to involve stakeholders in identifying objectives, problems, and strategies to overcome them. It involves communication planning, shared value engagement, early involvement, and clear rules of engagement. The clinicalNET community will promote innovations, publications, and applications supported by clinical protocols and trials. The project will also publish publications, fact sheets, and bulletins, and establish healthcare ethics and safety rules. Scientific meetings and innovation dissemination workshops will be organized to transfer research results to target groups.

## 6. CONCLUSION AND FUTURE DIRECTIONS

The proposed *clinicalNET* aims to foster innovation in the healthcare sector by fostering creativity and openness in research outcomes. By bridging the gap between academia and the healthcare business sector, it will promote risk management and research outcomes adoption as a market product. The clinicalNET will explore innovation in a broader context, focusing on underdeveloped opportunities in clinical trials, physician collaboration, and care delivery process improvement. Information and communication technologies (ICT) have the potential to address challenges faced by healthcare professionals, and clinical information management and collaboration will be thoroughly investigated in clinical trials. This approach will help bridge the gap between academia and healthcare businesses and promote successful innovation in the healthcare sector.

In conclusion, the *clinicalNET* community will focus on promotion of innovations, publications and application support of the clinical protocols and trials used.

Additional new members will join the clinicalNET during its life. Thus, the clinicalNET will achieve its mission, which is to provide a health holistic ecosystem approach that has the standards of an open, constantly evolving, as most as possible trusted, independent, co-working and co-creator of clinicalNET in providing healthcare information and management in evidence-based practice, and further support in research, publishing, education and consultation to researchers, organizations, and individuals in medical and healthcare. All these finally benefit patient care by promoting the highest standards of safety, quality, and -effectiveness in healthcare.

## Data Availability

All data produced in the present work are contained in the manuscript

